# Identifying Potential Drug Targets for Prostate Cancer from a Genetic Perspective: A Mendelian Randomization Study

**DOI:** 10.1101/2025.01.02.25319933

**Authors:** Jiaqing Huang, Zhiqiang Li, Yunqiao Lin

**Affiliations:** Ningde Municipal Hospital of Ningde Normal University, Fujian, China; Ningde Clinical Medical College of Fujian Medical University, Fujian, China

**Keywords:** Prostate cancer, Mendelian randomization, Drug targets, Molecular docking

## Abstract

**Background:** This study aimed to identify novel therapeutic targets for prostate cancer (PCa) utilizing expression quantitative trait loci (eQTL) data through Mendelian randomization (MR) analysis, while exploring the potential underlying mechanisms.

**Methods:** We employed MR analysis to evaluate the causal relationships between eQTLs and PCa. Cis-expression quantitative trait loci (cis-eQTL, exposure) data were obtained from the eQTLGen Consortium. GWAS data for prostate cancer were obtained from the UK Biobank Consortium and the FinnGen Consortium, with the UK Biobank Consortium data used for primary discovery and the FinnGen Consortium data used for replication and validation. Additionally, we conducted enrichment analysis, constructed protein interaction networks, predicted potential drugs, and performed molecular docking experiments to elucidate the functional significance and therapeutic reliability of identified targets.

**Results:** Our findings revealed that HOXA9, MPHOSPH6, SLC45A3, PBX2, and HLA-A are positively correlated with PCa risk, whereas PPARGC1A, FLOT2, TKT, CARNS1, GPBAR1, CSF1R, and TRAV21 showed negative associations. Molecular docking analysis demonstrated that GPBAR1 exhibited the highest binding affinity among the top five predicted drugs.

**Conclusions:** This study identified 12 promising drug targets for PCa through MR analysis. Therapeutics developed to target these genes are anticipated to enhance the success rate in clinical trials, thus enabling more efficient development of PCa treatments and potentially lowering overall drug development costs.

## 1 Introduction

Prostate cancer (PCa) ranks among the most prevalent malignancies in men, with its incidence rising significantly as the global population ages^[1]^. By 2030, it is projected that approximately 1.7 million new PCa cases will be diagnosed worldwide^[2]^. Current treatment modalities for advanced PCa encompass radical prostatectomy, radiotherapy, and androgen deprivation therapy (ADT)^[3]^. For patients with advanced disease, additional options such as immunotherapy—including sipuleucel-T and immune checkpoint inhibitors—alongside targeted therapies inhibiting poly (adenosine diphosphate-ribose) polymerase (PARP) (e.g., olaparib, niraparib, and rucaparib) should be considered^[4]^. PCa is often characterized as a “cold tumor,” exhibiting diminished antigen expression, impaired immune suppression, and low levels of immune infiltration. These attributes limit the efficacy of immunotherapies and contribute to resistance, cancer recurrence, and metastasis, often leading to unfavorable prognoses^[5]^.

In light of the substantial burden posed by advanced PCa, the pursuit of novel drug targets holds considerable clinical importance. Integrating genetic insights into drug development represents a vital strategy for optimizing this endeavor^[6]^. Recent studies indicate that drug targets backed by genetic validation are approximately twice as likely to succeed in reaching the market compared to those lacking such support^[7]^. Mendelian randomization (MR) serves as a statistical approach that utilizes genetic variants, specifically single nucleotide polymorphisms (SNPs), associated with exposures as instrumental variables to elucidate causal relationships between exposures and outcomes^[8]^. MR has emerged as an essential tool for the reassessment of existing drugs and the identification of new therapeutic targets^[9]^. Given that numerous circulating proteins are encoded by druggable target genes, they function as vital biological effectors for many approved or experimental medications. Consequently, insights derived from expression quantitative trait loci (eQTLs) linked to variations in drug-responsive gene expression can inform potential exposure factors for therapeutic targets^[10, 11]^.

## 2 Methods

The flowchart of this study is illustrated in Figure 1.

**Figure 1.**
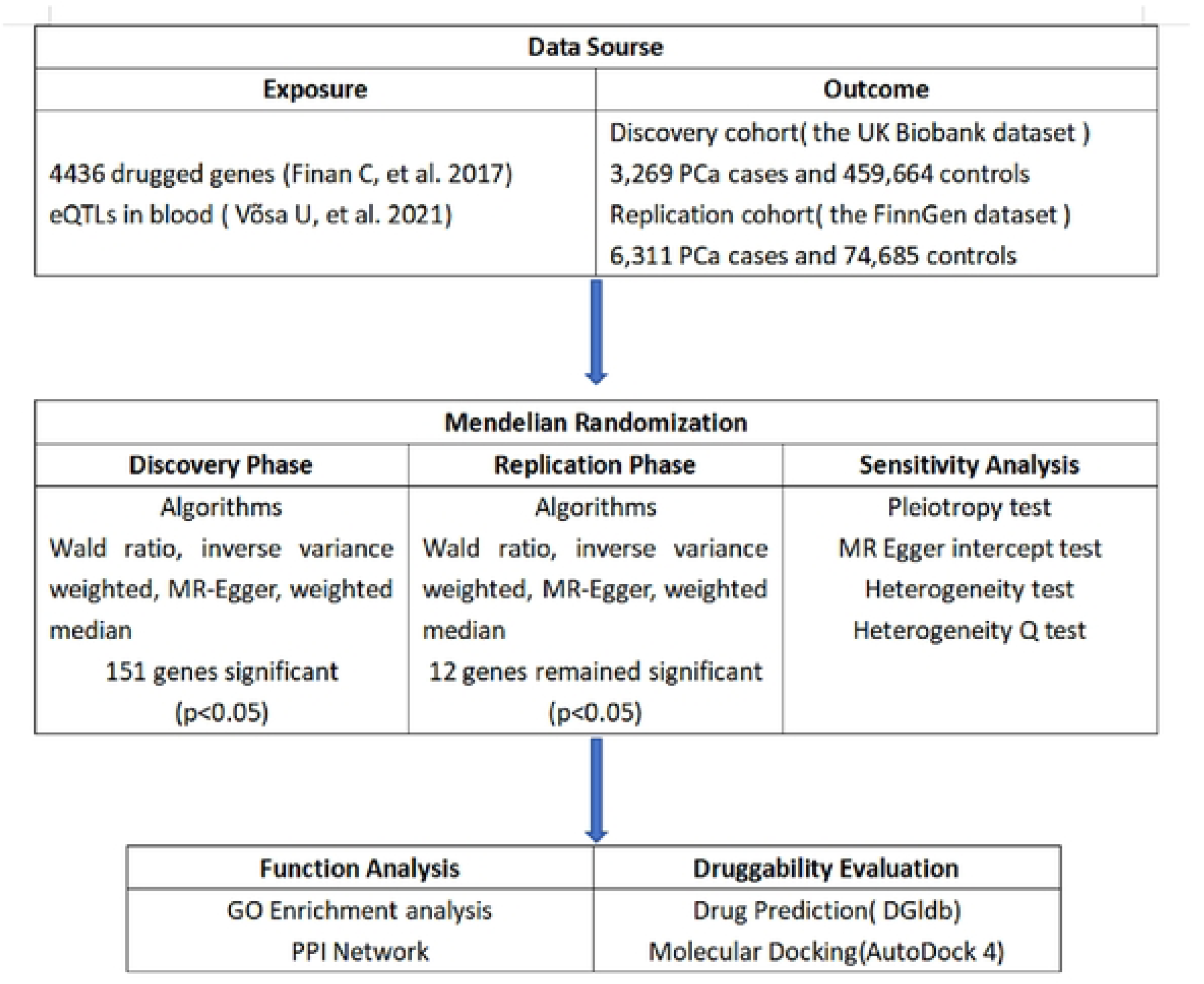
Overview of the study design

### 2.1 Exposure data

The eQTLGen dataset was obtained from the eQTLGen Consortium (https://eqtlgen.org/), comprising 16,987 genes and 31,684 cis-eQTLs. A comprehensive description of the dataset is provided in the original publication^[12]^. Druggable genes were identified from a compiled list of 4,463 genes associated with known drug targets and genome-wide association studies (GWAS)^[13]^.

### 2.2 Outcome data

GWAS data for prostate cancer were sourced from the UK Biobank, which includes 3,269 cases and 459,664 controls, and the FinnGen Consortium, which comprises 6,311 cases and 74,685 controls. The data from the UK Biobank were utilized for discovery, while the FinnGen data served for replication and validation.

### 2.3 Mendelian randomisation analysis

MR analysis was conducted using the R package “TwosampleMR”^[14]^ in which built-in functions (harmonise_data) were used to import and harmonize exposure and outcome data. MR satisfies three basic assumptions: (1) the genetic instrumental variable (IV) is directly related to the exposure; (2) the IV is unrelated to any undisclosed confounders that may affect the exposure-outcome relationship; and (3) the IV affects the outcome only through the exposure, thus ruling out any pleiotropic effects^[15]^. First, SNPs significantly associated with each exposure factor were screened out (p < 5×10-8). Next, SNPs that were physically distant (distance ≥10000 kb) and SNPs with low linkage disequilibrium (R^2^ < 0.001) were further excluded. We evaluated the strength of each SNP effect using the F statistic calculation formula: F = beta^2/se^2, where “beta” represents the size of the SNP effect and “se” represents the corresponding standard error. To ensure that the instrumental variables are strong enough to avoid bias, we used the F statistic to measure the power of IV. We defined weak IV as a case where the F statistic was less than 10, and excluded SNPs with an F statistic less than 10 to eliminate instruments with weak correlation^[16]^. In addition, Cochran’s Q test and MR-Egger intercept test can be used to assess heterogeneity and detect pleiotropy^[17]^. If the Cochran’s Q statistic test yields statistically significant results, a random-effects inverse variance weighted (IVW) model will be applied. Conversely, if the test does not demonstrate statistical significance, a fixed-effects model will be utilized.

### 2.4 Enrichment analysis

To investigate the functional characteristics and biological relevance of the identified potential therapeutic target genes, Gene Ontology (GO) and Kyoto Encyclopedia of Genes and Genomes (KEGG) enrichment studies were performed using R packages clusterProfiler and Pathview^[18]^. GO includes three terms: biological process (BP), molecular function (MF), and cellular component (CC). KEGG pathways provide metabolic pathway information.

### 2.5 Protein interaction network construction

To explore the relationships between proteins, we constructed a protein-protein interaction (PPI) network using STRING, setting a minimum confidence score of 0.4 for interactions while maintaining all other parameters at default levels^[19]^. Cytoscape (V3.9.1) was employed for the visualization of PPI results.

### 2.6 Drug candidate prediction

To assess the potential of target genes for drug development, we utilized the Drug Signatures Database (DGIdb) (https://old.dgidb.org/) to predict suitable drug candidates^[20]^. The identified target genes were uploaded to DSigDB for drug candidate prediction, evaluating their pharmacological activity.

### 2.7 Molecular docking

Molecular docking was employed to investigate the interactions between candidate drugs and their respective targets^[21]^. This technique simulates the interactions between receptors and ligands, predicting the position and orientation of ligands within the binding pocket while calculating the binding affinity of drug-target pairs. Structural data for the drugs were obtained from the PubChem compound database (https://pubchem.ncbi.nlm.nih.gov/), while protein structures were derived from the Protein Data Bank (PDB) (http://www.rcsb.org/). Molecular docking for the top five selected drugs and the proteins encoded by the corresponding target genes was performed using AutoDock (https://autodock.scripps.edu/)^[22]^. Prior to docking, water molecules and excess ligands were removed from the protein and ligand files, hydrogen atoms were added, and the grid box was adjusted to encompass all domains of each protein macromolecule. Unrestricted molecular motion was enabled to facilitate docking, and AutoDock was subsequently used to visualize the docking results.

## 3 Results

### 3.1 MR results in the discovery phase

The GWAS data from the UK Biobank database were downloaded, which contained 3,269 cases and 459,664 controls. Using the IVW method, we identified 151 genes whose expression was causally associated with prostate cancer risk (p < 0.05, Figure 2, Supplementary Table S1). The MR-Egger intercept test and Cochran Q test showed that these 151 genes did not have pleiotropy and heterogeneity (heterogeneity P > 0.05, pleiotropy P > 0.05; Supplementary Table S1).

**Figure 2.**
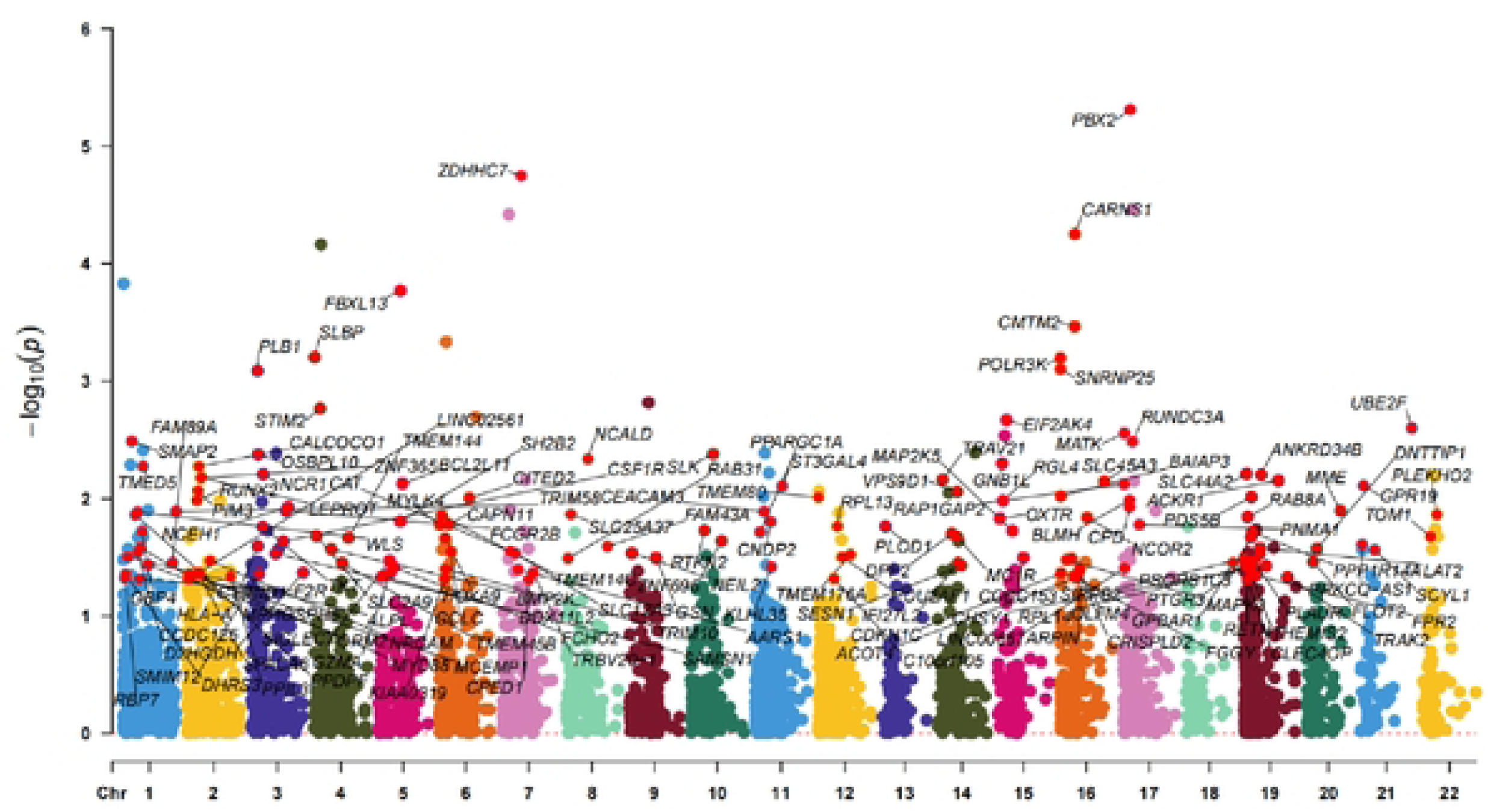
Manhattan plot of MR analysis in discovery phase. Significant genes were labelled.

### 3.2 MR results during the replication phase

This study used GWAS data from the FinnGen database, including 6311 cases and 74,685 controls. After IVW method validation of 151 genes, it was determined that the genetic predicted expression of 12 genes was causally related to prostate cancer risk (p < 0.05, Figure 3). The MR-Egger intercept test and Cochran Q test showed that these 12 genes did not have pleiotropy and heterogeneity (heterogeneity P > 0.05, pleiotropy P > 0.05, Supplementary Table S2).

**Figure 3.**
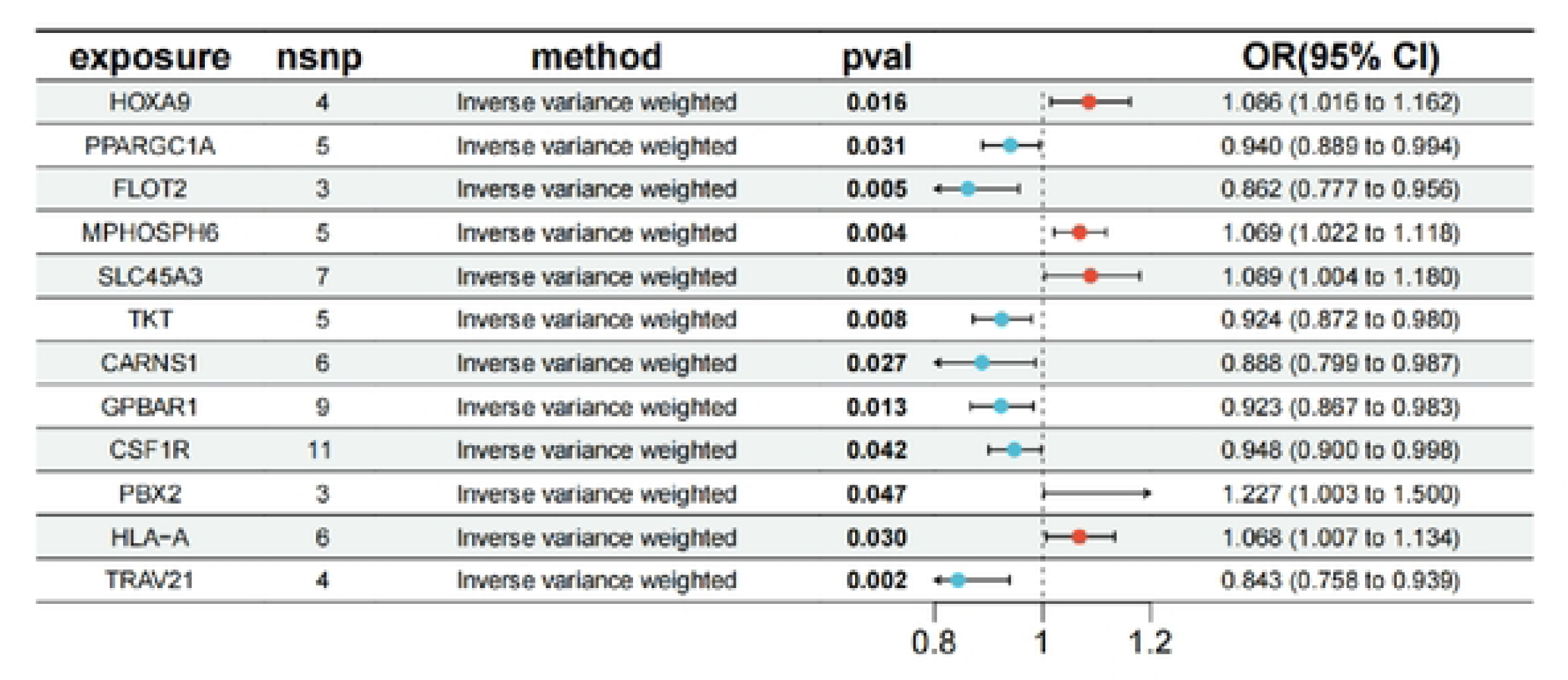
Forest plotsdisplaying the findings from the replication phase for 12 significant genes

### 3.3 Enrichment analysis

GO enrichment analysis was used to evaluate the role of genes of interest, and KEGG enrichment analysis clarified the relationship between genes and functional pathways^[23]^. From our analysis, it is evident that these genes are involved in various biological processes, including proximal/distal pattern formation, positive regulation of fatty acid metabolic process, positive regulation of glucose metabolic process, positive regulation of carbohydrate metabolic process, and regulation of fatty acid metabolic process. The cellular components include Golgi medial cisterna, uropod, cell trailing edge, and nuclear exosome (RNase complex). The molecular functions include T cell receptor binding, carbohydrate:monoatomic cation symporter activity, and acid-amino acid ligase activity (Figure 4a, b). The pathways of KEGG enrichment analysis were beta−Alanine metabolism, Histidine metabolism, Insulin signaling pathway, Pentose phosphate pathway, and Transcriptional misregulation in cancer (Figure 4c, d).

**Figure 4.**
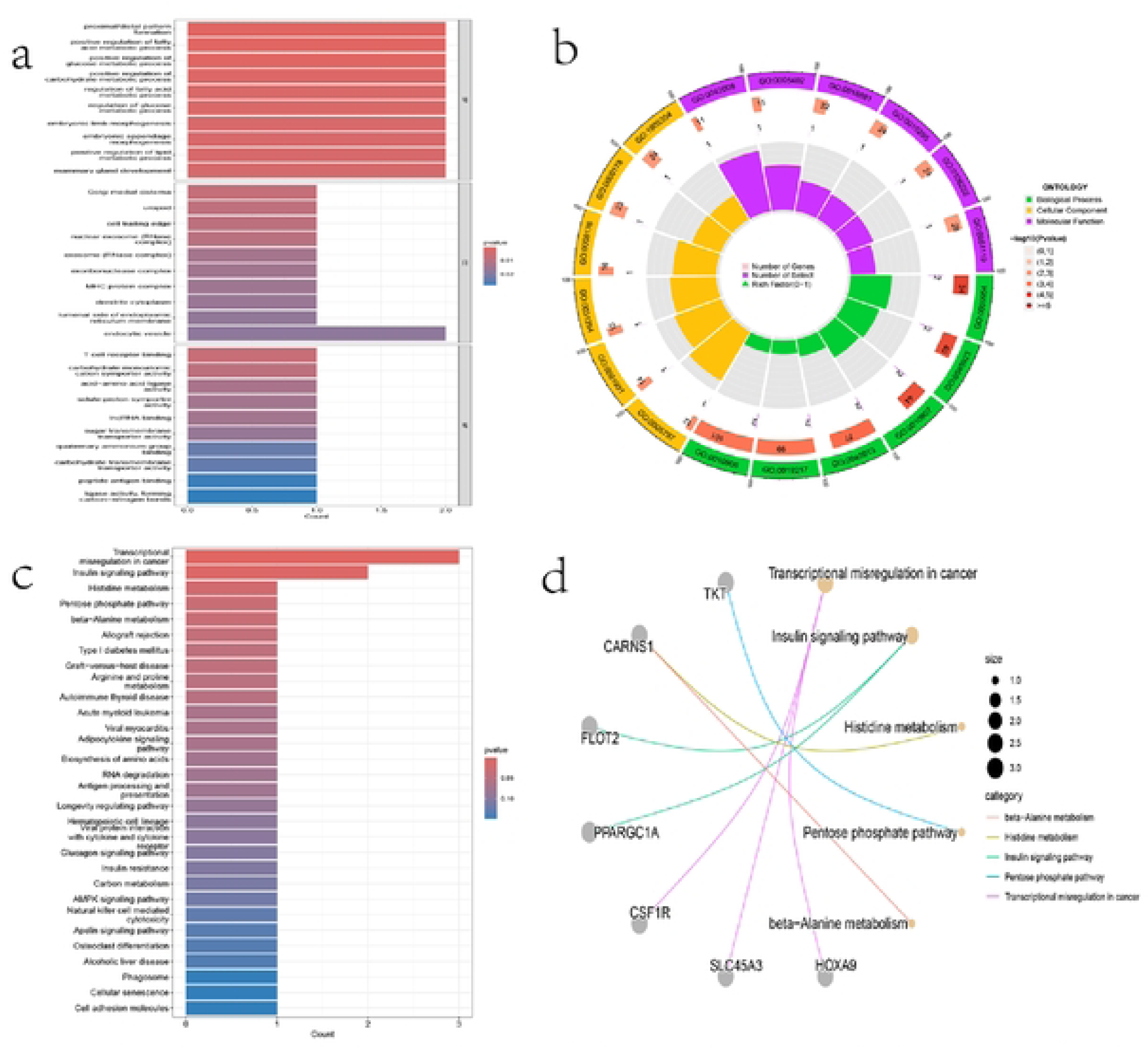
Functional enrichment analysis of twelve genes. (a)The barplot map of the GO of genes. (b) GO circlize map of genes. (c) The KEGG barplot map of genes. (d) The KEGG cnetplot map of genes.

### 3.4 PPI network

The 12 genes were loaded into the STRING (https://cn.string-db.org/) database to create a network, and the resulting file was imported into Cytoscape for visualization (Figure 5).

**Figure 5.**
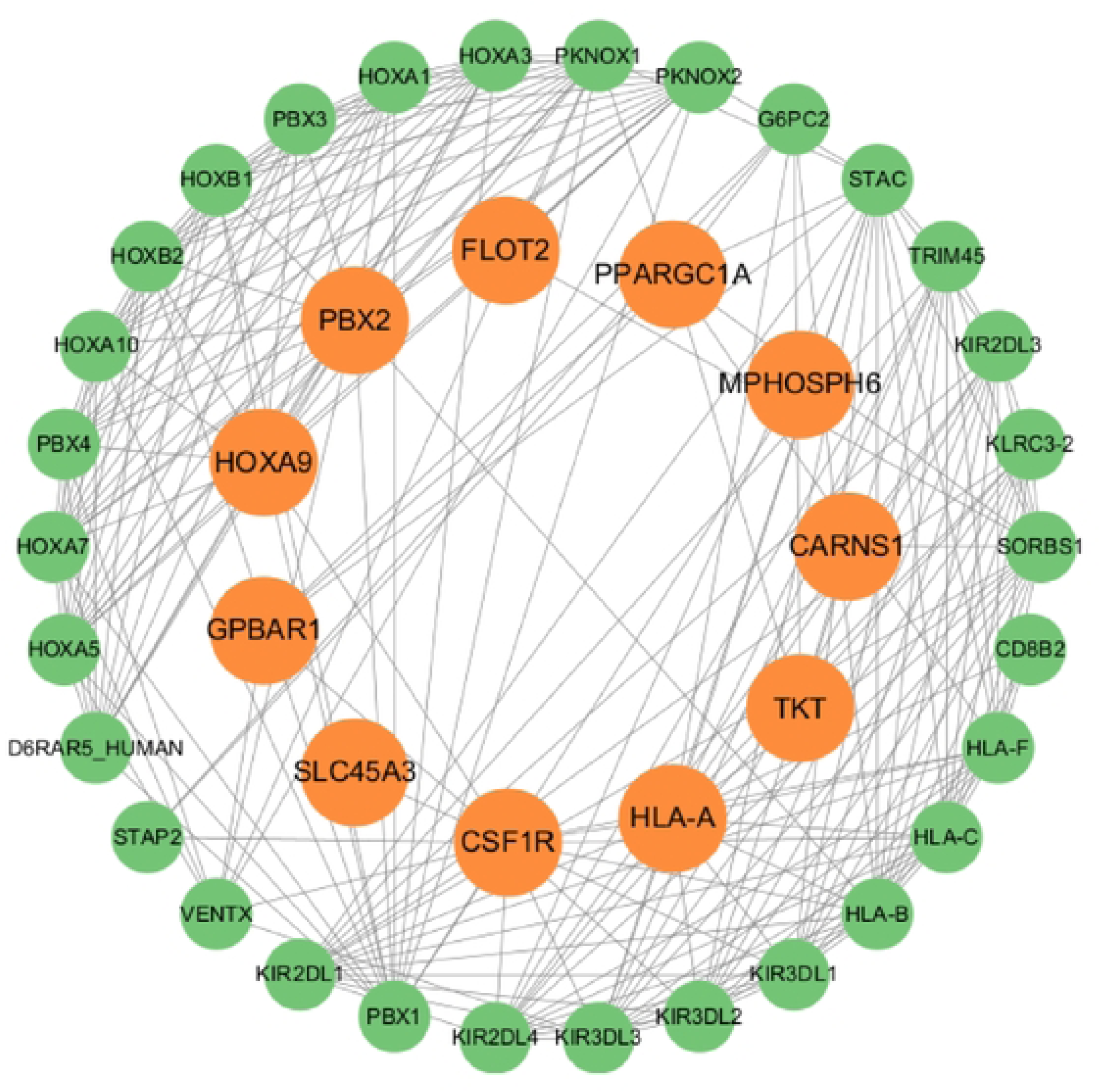
PPI network built with STRING

### 3.5 Drug candidate prediction

The DSigDB database was used to predict potentially effective intervention drugs. The top five candidate drugs were shown according to the adjusted p-values (Table 1).

**Table 1.**

| Drug names | zScore | pvalue | p.adjust | geneID |
| --- | --- | --- | --- | --- |
| Tauroursodeoxycholic acid | 15.6857 | 0.0001 | 0.0228 | PPARGC1A/GPBAR1 |
| Adehl | 7.0537 | 0.0024 | 0.0409 | PPARGC1A/GPBAR1 |
| GDP366 | 14.7140 | 0.0046 | 0.0409 | CSF1R |
| Decanoic acid | 14.7140 | 0.0046 | 0.0409 | PPARGC1A |
| GW612286X | 14.7140 | 0.0046 | 0.0409 | CSF1R |

Candidate drug predicted using DSigDB. The top five candidate drugs were shown.

### 3.6 Molecular docking

In order to evaluate the affinity of candidate drugs with their targets and understand the drugability of drug targets, molecular docking was performed in this study. AutoDock was used to perform molecular docking on the top five candidate drugs and the proteins encoded by the corresponding genes, and the binding energy of each interaction was calculated, resulting in a total of 7 protein-drug docking results (Figure 6, Table 2).

**Figure 6.**
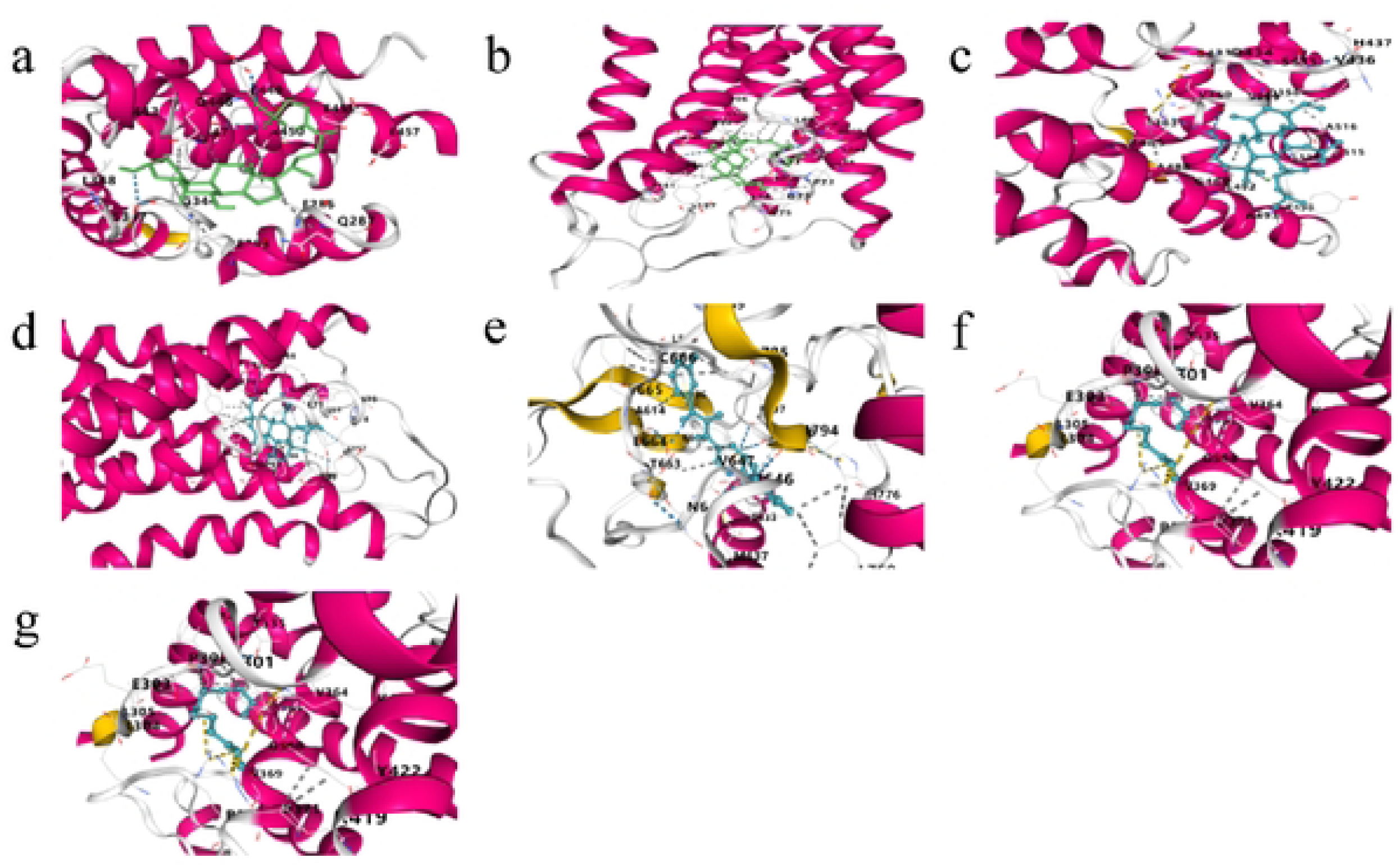
Docking results of available proteins small molecules. a PPARGClA docking Tauroursodeoxycholic acid, b GPBARl docking Tauroursodeoxycholic acid, c PPARGClA docking Adehl, d GPBARl docking Adehl, e CSFlR docked to GOP366, f PPARGClA docked to Oecanoic acid, g CSFlR docking GW612286X

**Table 2.**

| Target | Drug | CurPocket ID | Binding energy |
| --- | --- | --- | --- |
| PPARGC1A | Tauroursodeoxycholic acid | C3 | -7.2 |
| PPARGC1A | Adehl | C2 | -5.6 |
| PPARGC1A | Decanoic acid | C1 | -4.8 |
| GPBAR1 | Tauroursodeoxycholic acid | C4 | -11.1 |
| GPBAR1 | Adehl | C4 | -9.5 |
| CSF1R | GDP366 | C1 | -9.4 |
| CSF1R | GW612286X | C1 | -8.8 |

The lower the Binding Energy, the better the binding effect and the higher the affinity.

## 4 Discussion

Novel drug targets may potentially emerge from genes encoding specific proteins or RNAs. Pharmaceutical agents can interact with these molecules in a precise manner by modulating their expression or function. In this investigation, Mendelian randomization (MR) methodologies were employed to identify protein targets associated with prostate cancer, resulting in the discovery of 12 potential genes (HOXA9, MPHOSPH6, SLC45A3, PBX2, HLA-A, PPARGC1A, FLOT2, TKT, CARNS1, GPBAR1, CSF1R, and TRAV21). To further elucidate the biological functions of these protein targets, we conducted enrichment and protein-protein interaction (PPI) network analyses. Concurrently, we utilized the DSigDB database to predict efficacious intervention drugs for target genes and performed molecular docking experiments to validate the reliability of potential drug targets.

This study identified HOXA9, MPHOSPH6, SLC45A3, PBX2, and HLA-A as positively correlated with prostate cancer risk. HOXA9 is associated with endothelial cell (EC) proliferation and angiogenesis. Abnormal expression of HOXA9 can activate downstream signaling pathways, thereby creating a microenvironment conducive to tumor growth. Several studies have demonstrated that acute myeloid leukemia (AML), chronic myeloproliferative neoplasms (MPNs), and colon cancer are associated with overexpression of HOXA9, indicating a poor prognosis^[24–26]^. A previous study indicated that HOXA9 is involved in immune cell infiltration and in the recruitment and infiltration of dendritic cells, macrophages, and mast cells during the progression of prostate cancer^[27]^. This further identified HOXA9 as a risk gene for prostate cancer. MPHOSPH6 is an exosome-associated RNA-binding protein involved in 5.8S rRNA maturation, and its actions affect the cell cycle and ovarian development^[28]^.SLC45A3 encodes the solute carrier family 45, member 3 protein, also known as prostate cancer-associated protein 6, which is primarily involved in the positive regulation of small molecule metabolic processes and the regulation of oligodendrocyte differentiation^[29]^. Additionally, SLC45A3 is also considered to be related to prostate-specific antigen serum concentration and prostate cancer^[30]^. PBX2 is a transcriptional activator that binds to the TLX1 promoter, thereby inducing the execution of various cellular functions, such as anti-apoptosis and inhibition of cell differentiation and proliferation^[31]^. Previous studies have found that high expression of PBX2 in various cancer tissues, including lung, stomach, and esophagus, is associated with poor prognosis^[32]^. HLA-A is a homolog of the human leukocyte antigen (HLA) class I heavy chain and plays a central role in the immune system by presenting peptides in the lumen of the endoplasmic reticulum and can be recognized by cytotoxic T cells^[33]^. In addition, previous studies have reported a positive correlation between HLA-A and the progression of prostate cancer, which is consistent with our results^[34]^.

PPARGC1A, FLOT2, TKT, CARNS1, GPBAR1, CSF1R, and TRAV21 were negatively correlated with PCa risk. The protein encoded by PPARGC1A is a transcriptional coactivator that regulates genes involved in energy metabolism. PPARGC1A has been reported to affect the prognosis of anaplastic thyroid cancer by inhibiting immune cell infiltration^[35]^. In addition, a LASSO analysis revealed that PPARGC1A has a protective role in prostate cancer^[36]^. FLOT2 encodes an integral membrane protein involved in vesicle trafficking and capable of interacting with signaling molecules such as kinases, G proteins, and adhesion molecule receptors^[37]^. FLOT2 has been shown to be involved in the development of cancer, such as playing an important role in the metastasis of nasopharyngeal carcinoma cells, the occurrence and prognosis of colorectal cancer. In addition, studies have shown that FLOT2 can inhibit the growth of cervical cancer in vivo^[38]^. TKT encodes a thiamine-dependent enzyme that directs excess sugar phosphate into glycolysis in the pentose phosphate pathway and plays an essential role in nucleic acid synthesis and cell division^[39]^. Previous studies have also confirmed that TKT is transported into the cell nucleus, thereby inhibiting the activity of the FXR promoter, affecting hepatic bile acid metabolism, and promoting the occurrence of liver cancer^[40]^. CARNS1 is a protein containing an ATP-grip domain that catalyzes the synthesis of carnosine and homoeosin^[41]^. Carnosine inhibits glycolytic energy metabolism in human malignancies, which is essential for cancer cells. Some scholars also pointed out that carnosine treatment can reduce the proliferation of gastric cancer cells and promote cell apoptosis by activating Akt/mTOR signaling and inducing cell cycle arrest^[42]^. GPBAR1 encodes a bile acid G protein-coupled receptor (GPCR). Studies have shown that GPBAR1 can reduce the inflammatory response caused by liver ischemia-reperfusion injury and inhibit hepatocyte apoptosis^[43]^. Our study found that GPBAR1 was negatively correlated with prostate cancer risk, but another study obtained inconsistent results^[44]^. CSF1R protein encodes a receptor that exacerbates the overall tumor microenvironment (TME) by persistent oxidative stress in castration-resistant prostate cancer (CRPC) cells, and thus targeting the TME by inhibiting colony-stimulating factor 1 receptor (CSF1R) is a very promising CRPC treatment approach^[45]^. DeNardo et al. also highlighted the importance of CSF1/CSF1R signaling in the recruitment of tumor-associated macrophages (TAMs) in breast cancer and further suggested a role for CSF1R in improving therapy^[46]^. TRAV21 encodes a T cell receptor that recognizes foreign antigens, and the key function of NKT cells lies in the interaction between their TCR and antigen complexes, which may have an inhibitory effect on tumor growth^[47]^.

A strength of the current study is the use of UK and Finnish genetic databases, one for the discovery cohort and the other for the replication cohort, significantly reducing the risk of false-positive results in identifying potential drug targets for prostate cancer. To further validate the dependability of these targets, the study also employed molecular docking was used to assess the binding affinity of target genes and possible medications. However, this study has some limitations. First, it lacks consideration of factors other than genetic, such as environmental and lifestyle factors. At the same time, for potential drug targets identified by MR and analytical docking methods, further experimental validation and rigorous clinical trials are needed to evaluate their safety and efficacy.

## 5 Conclusions

In summary, this study demonstrated that HOXA9, MPHOSPH6, SLC45A3, PBX2, HLA-A, PPARGC1A, FLOT2, TKT, CARNS1, GPBAR1, CSF1R, and TRAV21 are associated with prostate cancer and provides important insights into targeted therapy for this disease. Further research and clinical trials on these genes and drugs are necessary.

## Data Availability

https://gwas.mrcieu.ac.uk/datasets/ukb-b-13348https://www.finngen.fi/en

https://www.finngen.fi/enhttps://gwas.mrcieu.ac.uk/datasets/ukb-b-13348

## Abbreviations

PCa: Prostate cancer
GWAS: Genome-wide association studies
SNP: Single Nucleotide Polymorphism
MR: Mendelian randomization
eQTL: Expression quantitative trait loci
cis-eQTL: Cis-expression quantitative trait loci
IVW: Inverse variance weighted
GO: Gene Ontology
KEGG: Kyoto Encyclopedia of Genes and Genomes
PPI: Protein–protein interaction.

## Availability of Data and Materials

The datasets analysed during the current study are available in the in the following repositories: eQTLs data were obtained from eQTLGen Consortium (https://eqtlgen.org/); PCa discovery data was downloaded on GWAS Catalog (https://gwas.mrcieu.ac.uk/datasets/ukb-b-13348); PCa replication data was from FinnGen Release 10 (https://www.finngen.fi/en).

## Author Contributions

Yunqiao Lin developed a major research plan. Jiaqing Huang and Zhiqiang Li analyze data, draw charts and write manuscripts. Yunqiao Lin helped collect data and references. All authors contributed to the article and approved the submitted version.

## Ethics Approval and Consent to Participate

The ethical review and approval for the original studies are available in the respective publications. This Mendelian randomization (MR) study utilized summary-level statistics only, ensuring that no identifiable private information from the GWAS datasets was accessed or included in the analysis.

## Acknowledgment

We thank the eQTLGen consortium, the FinnGen team and other researchers and participants for providing publicly available data for this analysis.

## References

[1] Sung H, Ferlay J, Siegel R L, et al. Global Cancer Statistics 2020: Globocan Estimates of Incidence and Mortality Worldwide for 36 Cancers in 185 Countries [J]. Ca Cancer J Clin, 2021, 71(3): 209–49.

[2] Ferlay J, Shin H R, Bray F, et al. Estimates of worldwide burden of cancer in 2008: Globocan 2008 [J]. Int J Cancer, 2010, 127(12): 2893–917.

[3] Eastham J A, Auffenberg G B, Barocas D A, et al. Clinically Localized Prostate Cancer: AUA/Astro Guideline, Part I: Introduction, Risk Assessment, Staging, and Risk-Based Management [J]. J Urol, 2022, 208(1): 10–8.

[4] Cha H R, Lee J H, Ponnazhagan S. Revisiting Immunotherapy: A Focus on Prostate Cancer [J]. Cancer Res, 2020, 80(8): 1615–23.

[5] Ebelt K, Babaryka G, Frankenberger B, et al. Prostate cancer lesions are surrounded by FOXP3+, PD-1+ and B7-H1+ lymphocyte clusters [J]. Eur J Cancer, 2009, 45(9): 1664–72.

[6] Nelson M R, Tipney H, Painter J L, et al. The support of human genetic evidence for approved drug indications [J]. Nat Genet, 2015, 47(8): 856–60.

[7] King E A, Davis J W, Degner J F. Are drug targets with genetic support twice as likely to be approved? Revised estimates of the impact of genetic support for drug mechanisms on the probability of drug approval [J]. PLoS Genet, 2019, 15(12): e1008489.

[8] Emdin C A, Khera A V, Kathiresan S. Mendelian Randomization [J]. JAMA, 2017, 318(19): 1925–6.

[9] Storm C S, Kia D A, Almramhi M M, et al. Finding genetically-supported drug targets for Parkinson’s disease using Mendelian randomization of the druggable genome [J]. Nat Commun, 2021, 12(1): 7342.

[10] Reay W R, Cairns M J. Advancing the use of genome-wide association studies for drug repurposing [J]. Nat Rev Genet, 2021, 22(10): 658–71.

[11] Schmidt A F, Finan C, Gordillo-Maranon M, et al. Genetic drug target validation using Mendelian randomisation [J]. Nat Commun, 2020, 11(1): 3255.

[12] Vosa U, Claringbould A, Westra H J, et al. Large-scale cis- and trans-eqtl analyses identify thousands of genetic loci and polygenic scores that regulate blood gene expression [J]. Nat Genet, 2021, 53(9): 1300–10.

[13] Finan C, Gaulton A, Kruger F A, et al. The druggable genome and support for target identification and validation in drug development [J]. Sci Transl Med, 2017, 9(383).

[14] Hemani G, Zheng J, Elsworth B, et al. The MR-Base platform supports systematic causal inference across the human phenome [J]. Elife, 2018, 7.

[15] Davies N M, Holmes M V, Davey Smith G. Reading Mendelian randomisation studies: a guide, glossary, and checklist for clinicians [J]. BMJ, 2018, 362: k601.

[16] Burgess S, Thompson S G, Collaboration C C G. Avoiding bias from weak instruments in Mendelian randomization studies [J]. Int J Epidemiol, 2011, 40(3): 755–64.

[17] Greco M F, Minelli C, Sheehan N A, et al. Detecting pleiotropy in Mendelian randomisation studies with summary data and a continuous outcome [J]. Stat Med, 2015, 34(21): 2926–40.

[18] Luo W, Brouwer C. Pathview: an R/Bioconductor package for pathway-based data integration and visualization [J]. Bioinformatics, 2013, 29(14): 1830–1.

[19] Szklarczyk D, Kirsch R, Koutrouli M, et al. The String database in 2023: protein-protein association networks and functional enrichment analyses for any sequenced genome of interest [J]. Nucleic Acids Res, 2023, 51(D1): D638–D46.

[20] Yoo M, Shin J, Kim J, et al. DSigDB: drug signatures database for gene set analysis [J]. Bioinformatics, 2015, 31(18): 3069–71.

[21] Morris G M, Huey R, Olson A J. Using AutoDock for ligand-receptor docking [J]. Curr Protoc Bioinformatics, 2008, Chapter 8: Unit 8 14.

[22] Kim S, Chen J, Cheng T, et al. PubChem in 2021: new data content and improved web interfaces [J]. Nucleic Acids Res, 2021, 49(D1): D1388–D95.

[23] Chen L, Zhang Y H, Lu G, et al. Analysis of cancer-related lncRNAs using gene ontology and Kegg pathways [J]. Artif Intell Med, 2017, 76: 27–36.

[24] Talarmain L, Clarke M A, Shorthouse D, et al. HOXA9 has the hallmarks of a biological switch with implications in blood cancers [J]. Nat Commun, 2022, 13(1): 5829.

[25] Lambert M, Alioui M, Jambon S, et al. Direct and Indirect Targeting of HOXA9 Transcription Factor in Acute Myeloid Leukemia [J]. Cancers (Basel), 2019, 11(6).

[26] Bhatlekar S, Viswanathan V, Fields J Z, et al. Overexpression of HOXA4 and HOXA9 genes promotes self-renewal and contributes to colon cancer stem cell overpopulation [J]. J Cell Physiol, 2018, 233(2): 727–35.

[27] Song Y P, Xian P, Luo H, et al. Comprehensive Landscape of HOXA2, HOXA9, and HOXA10 as Potential Biomarkers for Predicting Progression and Prognosis in Prostate Cancer [J]. J Immunol Res, 2022, 2022: 5740971.

[28] Schilders G, Raijmakers R, Raats J M, et al. MPP6 is an exosome-associated RNA-binding protein involved in 5.8S rrna maturation [J]. Nucleic Acids Res, 2005, 33(21): 6795–804.

[29] Walker M G, Volkmuth W, Sprinzak E, et al. Prediction of gene function by genome-scale expression analysis: prostate cancer-associated genes [J]. Genome Res, 1999, 9(12): 1198–203.

[30] Pin E, Henjes F, Hong M G, et al. Identification of a Novel Autoimmune Peptide Epitope of Prostein in Prostate Cancer [J]. J Proteome Res, 2017, 16(1): 204–16.

[31] Qiu Y, Wang Z L, Jin S Q, et al. Expression level of pre-B-cell leukemia transcription factor 2 (PBX2) as a prognostic marker for gingival squamous cell carcinoma [J]. J Zhejiang Univ Sci B, 2012, 13(3): 168–75.

[32] Qiu Y, Morii E, Tomita Y, et al. Prognostic significance of pre B cell leukemia transcription factor 2 (PBX2) expression in non-small cell lung carcinoma [J]. Cancer Sci, 2009, 100(7): 1198–209.

[33] Manca M A, Simula E R, Cossu D, et al. Association of HLA-A*11:01, -A*24:02, and -B*18:01 with Prostate Cancer Risk: A Case-Control Study [J]. Int J Mol Sci, 2023, 24(20).

[34] Stokidis S, Fortis S P, Kogionou P, et al. Hla Class I Allele Expression and Clinical Outcome in De Novo Metastatic Prostate Cancer [J]. Cancers (Basel), 2020, 12(6).

[35] Huang Y, Ling J, Chang A, et al. Identification of an immune-related key gene, PPARGC1A, in the development of anaplastic thyroid carcinoma: in-silico study and in-vitro evaluation [J]. Minerva Endocrinol (Torino), 2022, 47(2): 150–9.

[36] Lv D, Wu X, Chen X, et al. A novel immune-related gene-based prognostic signature to predict biochemical recurrence in patients with prostate cancer after radical prostatectomy [J]. Cancer Immunol Immunother, 2021, 70(12): 3587–602.

[37] Kwiatkowska K, Matveichuk O V, Fronk J, et al. Flotillins: At the Intersection of Protein S-Palmitoylation and Lipid-Mediated Signaling [J]. Int J Mol Sci, 2020, 21(7).

[38] Li Y, Dou S. FLOT2 Promotes the Proliferation and Epithelial-mesenchymal Transition of Cervical Cancer by Activating the MEK/ERK1/2 Pathway [J]. Balkan Med J, 2022, 39(4): 267–74.

[39] Li M, Zhao X, Yong H, et al. Transketolase promotes colorectal cancer metastasis through regulating Akt phosphorylation [J]. Cell Death Dis, 2022, 13(2): 99.

[40] Li M, Zhang X, Lu Y, et al. The nuclear translocation of transketolase inhibits the farnesoid receptor expression by promoting the binding of HDAC3 to Fxr promoter in hepatocellular carcinoma cell lines [J]. Cell Death Dis, 2020, 11(1): 31.

[41] Drozak J, Veiga-Da-Cunha M, Vertommen D, et al. Molecular identification of carnosine synthase as ATP-grasp domain-containing protein 1 (ATPGD1) [J]. J Biol Chem, 2010, 285(13): 9346–56.

[42] Shen Y, Yang J, Li J, et al. Carnosine inhibits the proliferation of human gastric cancer SGC-7901 cells through both of the mitochondrial respiration and glycolysis pathways [J]. PLoS One, 2014, 9(8): e104632.

[43] Yang H, Zhou H, Zhuang L, et al. Plasma membrane-bound G protein-coupled bile acid receptor attenuates liver ischemia/reperfusion injury via the inhibition of toll-like receptor 4 signaling in mice [J]. Liver Transpl, 2017, 23(1): 63–74.

[44] Cai J, Zhao J, Gao P, et al. Patchouli alcohol inhibits GPBAR1-mediated cell proliferation, apoptosis, migration, and invasion in prostate cancer [J]. Transl Androl Urol, 2022, 11(11): 1555–67.

[45] Xu J, Escamilla J, Mok S, et al. CSF1R signaling blockade stanches tumor-infiltrating myeloid cells and improves the efficacy of radiotherapy in prostate cancer [J]. Cancer Res, 2013, 73(9): 2782–94.

[46] Denardo D G, Brennan D J, Rexhepaj E, et al. Leukocyte complexity predicts breast cancer survival and functionally regulates response to chemotherapy [J]. Cancer Discov, 2011, 1(1): 54–67.

[47] Paul S, Chhatar S, Mishra A, et al. Natural killer T cell activation increases iNOS(+)CD206(-) M1 macrophage and controls the growth of solid tumor [J]. J Immunother Cancer, 2019, 7(1): 208.

